# Effects of Whole-Body Vibration Training on Muscle Strength and Flexibility: A Meta-Analysis with Subgroup Analyses Based on Population and Vibration Parameters

**DOI:** 10.1101/2025.11.10.25339946

**Authors:** Jingui Huang, ke Sun

**Affiliations:** Yunnan Normal University, China; Yunnan University of Finance and Economics, China

**Keywords:** Whole-body vibration training, Muscle strength, Flexibility, Meta-analysis, Vibration parameters

## Abstract

This study conducted a meta-analysis to systematically evaluate the effects of whole-body vibration training on muscle strength and flexibility in different athletic populations, as well as to examine the moderating effects of factors such as vibration frequency, training duration, amplitude, and population type on the outcomes, aiming to provide a basis for the scientific application of vibration training.Randomized controlled trials related to whole-body vibration training were retrieved from databases including PubMed, Web of Science, CNKI, VIP, Wanfang, EBSCO, and Cochrane. Literature was screened based on the PICOS principle, and Meta-analysis and subgroup analyses were conducted using Review Manager 5.4 to evaluate the overall effects of vibration training on strength and flexibility. A total of 15 studies involving 491 participants were included, encompassing healthy young adults, the elderly, athletes, special female populations, and special male populations.The meta-analysis results showed that the overall effect size of whole-body vibration training on strength was moderate (SMD=0.67, 95% CI [0.42, 0.93]), while the overall effect size on flexibility was small (SMD=0.20, 95% CI [-0.05, 0.45]).Subgroup analyses indicated that vibration training yielded the most pronounced strength improvement in special female populations, whereas its effect on healthy young adults was relatively limited (SMD=0.13). In terms of vibration parameters, medium frequency (20-40 Hz) and medium amplitude (2-4 mm) yielded optimal effects. Medium- to long-term training durations (8-16 weeks and >16 weeks) were superior to short-term interventions (≤ 8 weeks). For flexibility, the overall effect remained non-significant (SMD=0.20).In conclusion, whole-body vibration training demonstrates a moderate effect on improving muscle strength, with particularly significant benefits observed in special female populations and the elderly. However, its effect on flexibility enhancement remains limited. Vibration frequency, amplitude, and training duration are key factors influencing the training outcomes. Future research should focus on further optimizing vibration parameters and conducting empirical studies on personalized training protocols.

## 1. Introduction

With the increasingly fierce competition in competitive sports and the gro wing demand for scientific approaches in public fitness, exploring efficient and innovative training methods has become a crucial pathway for enhancing athletic performance (Bompa & Haff, 2009). As fundamental components of physical fitness (NSCA, 2016), strength and flexibility serve not only as essential safe guards against sports injuries (Thacker et al., 2004) but also as key determinants of movement execution quality and competitive performance levels (Young & Behm, 2002).While traditional resistance training and static stretching can effectively develop strength and flexibility, their relatively monotonous training modalities, tendency to plateau, and limitations in specific populations have prompted researchers to seek more optimal training solutions (Lim JS et al., 2013).

In recent years, whole-body vibration (WBV) has gained increasing attention in sports science as a novel, low-load exercise modality that minimally interferes with regular training (He Limin et al., 2018). WBV training involves standing on a vibrating platform where generated vibrations stimulate muscle spindles and trigger alpha motor neuron reflexes. The vertical acceleration is transmitted to muscles and bones, thereby inducing plyometric contractions in the muscles (Lai CL et al., 2014).Although the beneficial effects of aerobic and resistance exercise on physical health have been well-documented (Manthou E et al., 2015), it is often challenging to engage patients with limited active motor function in voluntary training. Whole-body vibration (WBV) training effectively bridges this gap.

Current research has consistently demonstrated the efficacy of such training. For instance, vibration intervention has shown significant effects in improving hamstring flexibility and sit-and-reach performance (Fagnani et al., 2006), while vibration training at specific frequencies has been proven to substantially enhance lower limb explosive power (Rønnestad, 2009).Nevertheless, the efficacy of vibration training remains debated in the academic literature(Marin & Rhea, 2010). However, other studies have reached contradictory conclusions. This state of conflicting evidence indicates that simply examining whether vibration training is effective is no longer sufficient to advance the field; a more in-depth exploration of the key factors influencing training outcomes is required.In-depth analysis reveals that the inconsistencies in training outcomes primarily stem from the multi-parametric nature of vibration training itself. Among these parameters, vibration frequency and training duration have been identified as critical moderating variables (Cardinale & Wakeling, 2005; Pollock et al., 2010).Regarding vibration frequency parameters, existing studies have utilized a frequency spectrum ranging from 20 Hz to over 50 Hz, where different frequencies may elicit distinctly different training effects (Cardinale & Wakeling, 2005). In terms of training duration, short-term interventions (typically ≤ 8 weeks) primarily induce neural adaptations and transient improvements in flexibility, whereas medium- to long-term training (usually > 8 weeks) is more likely to produce sustained structural and functional muscular adaptations (Pollock et al., 2010).Heterogeneity among participant populations is also a significant factor contributing to divergent research findings, as different groups exhibit inherent differences in neuromuscular system characteristics, training background, and recovery capacity (Haff & Triplett, 2016/2019). For well-trained athletes, vibration training can serve as a novel stimulus to overcome training plateaus; whereas for novice trainees, its effects may be overshadowed by improvements derived from foundational training (Cardinale & Wakeling, 2005).

Most existing reviews have failed to adequately differentiate the response characteristics of diverse populations to vibration stimuli, thereby limiting the generalizability and practical value of their conclusions. This study employs a meta-analytic approach to systematically evaluate the effects of vibration interventions on strength and flexibility across different populations. Furthermore, subgroup analyses will be conducted to examine the intervention effects under various vibration parameters and training durations, aiming to provide scientific evidence for optimizing vibration intervention strategies.

## 2. Research Methods

The design and reporting of this systematic review and meta-analysis complied with the PRISMA (Preferred Reporting Items for Systematic Reviews and Meta-Analyses) guidelines (Liberati A et al., 2009).This systematic review and meta-analysis were conducted according to the Preferred Reporting Items for Systematic Reviews and Meta-Analyses statement guidelines. The study protocol was registered in the International Prospective Register of Systematic Reviews ID:CRD420251173639.

### Literature Search

A systematic literature search was performed across seven electronic databases—PubMed, Web of Science, CNKI, VIP, Wanfang, EBSCO, and Cochrane—for studies published between 1994 and October 13, 2025, investigating the effects of vibration interventions on strength and flexibility. The final search was performed on October 13, 2025.

Search Strategy:(Vibratory training OR Whole-body vibration OR Whole-body vibration training OR Vibration platform training)AND(Muscles trength OR Flexibility)AND(athletes OR Elderly people OR Teenagers)AND(Effect OR Impact OR Influence) 中文检索式:(振动训练 OR 全身振动 OR 全身振动训练 OR 振动平台训练)AND(力量 OR 柔韧性)AND(运动员 OR 老年人OR 青少年)AND(效果 OR 影响 OR 干预效果)AND(随机对照试验 OR 临床试验)

### Literature Inclusion and Exclusion Criteria

The study selection followed the PICOS framework, with inclusion criteria established based on five key components: Participants, Intervention, Comparison, Outcomes, and Study Design.

### Inclusion Criteria

1) Studies with a clearly defined whole-body vibration (WBV) intervention protocol in the experimental group. 2) Studies that provided sample size data and reported outcomes related to training modalities combined with WBV or other physiological parameters measured in participants of any age or sex. 3) Studies that included a control group receiving either conventional training or no training.

### Exclusion Criteria

1)Studies with outcome measures unrelated to “strength/flexibility” or employing non-standardized assessment methods.2)Participants who had undergone any vibration intervention prior to the trial.3)Study designs that preclude causal inference, or where essential data are missing and effect sizes cannot be calculated.

### Literature Screening and Data Extraction

First, all retrieved records were imported into Zotero for deduplication. Two investigators (the first and second authors of this paper) independently screened the literature according to the predetermined inclusion and exclusion criteria. The initial screening was performed by reviewing titles and abstracts. Any discrepancies or disagreements regarding the eligibility of specific studies were resolved through discussion with a third researcher to reach a consensus. Finally, relevant data were extracted from all studies that met the inclusion criteria.

The following data were systematically extracted from the included studies: authors, publication year, sample sizes of the experimental and control groups, participant age, intervention duration, intervention type, sport category, and outcome measures (e.g., strength, flexibility, or other relevant biological indicators).

### Quality Assessment

Two independent reviewers conducted the risk of bias assessment for the included studies using Review Manager 5.4 (software provided by the Cochrane Collaboration). They cross-verified their assessments, and studies with low methodological quality were excluded. The quality assessment results were categorized as “Low Risk,” “Unclear,” or “High Risk.” The evaluation criteria included randomization, allocation concealment, blinding, incomplete outcome data, selective reporting, and other potential sources of bias.

### Statistical Analysis

Data analysis was performed using Review Manager 5.4 software. Since all included data were continuous variables, the standardized mean difference (SMD) was used to calculate the pooled effect size, along with its 95% confidence interval. The interpretation of effect sizes followed Cohen’s criteria: SMD = 0.2 (small effect), 0.5 (medium effect), and ≥ 0.8 (large effect). It is important to note that in this study, a positive SMD value indicates an improvement in strength or flexibility, with higher SMD values representing stronger intervention effects. Given that some of the included studies exhibited baseline differences, the effect sizes were calculated using pre- to post-intervention change scores.The magnitude of heterogeneity among the included studies was assessed using the I² statistic, which ranges from 0% to 100%. A higher I^2^ value indicates greater heterogeneity. According to the Cochrane Handbook for Systematic Reviews (Higgins J et al., 2019), I² values are interpreted as follows: 0%–40% (might not be important), 30%–60% (may represent moderate heterogeneity), 50%–90% (may represent substantial heterogeneity), and 75%– 100% (considerable heterogeneity). To address heterogeneity, subgroup analyses were performed. By examining effect size differences across various subgroups, this approach helped determine whether the overall effect remained consistent across different populations and identify potential sources of heterogeneity in the research findings.

To further investigate the potential influence of different training parameters on the effects of vibration training, univariable and multivariable regression analyses were performed following the subgroup analyses (Weisberg, 2014). Specifically, univariable regression analyses were first conducted using frequency, amplitude, and cycle as independent variables, respectively, to identify variables significantly or marginally associated with the effect size. Subsequently, variables showing significance or a trend toward significance were simultaneously included in a multivariable regression model to further evaluate their independent effects. The models utilized restricted maximum likelihood (REML) estimation for random-effects variance components (Harville, 1977). Model fit was compared using the Akaike Information Criterion (AIC) (Burnham & Anderson, 2002) and the proportion of heterogeneity explained (R ²) to assess the goodness-of-fit across different models (Rights & Sterba, 2019).

## 3 Results and Analysis

### 3.1 Literature Search Results

A total of 642 articles related to the research topic were retrieved from seven databases. After importing the records into Zotero and removing 376 duplicates, the remaining articles were screened by title, abstract, and full-text review according to the predefined inclusion and exclusion criteria. Ultimately, 15 studies were included, comprising 14 international publications and 1 Chinese study, as illustrated in Figure 1.

**Figure 1.**
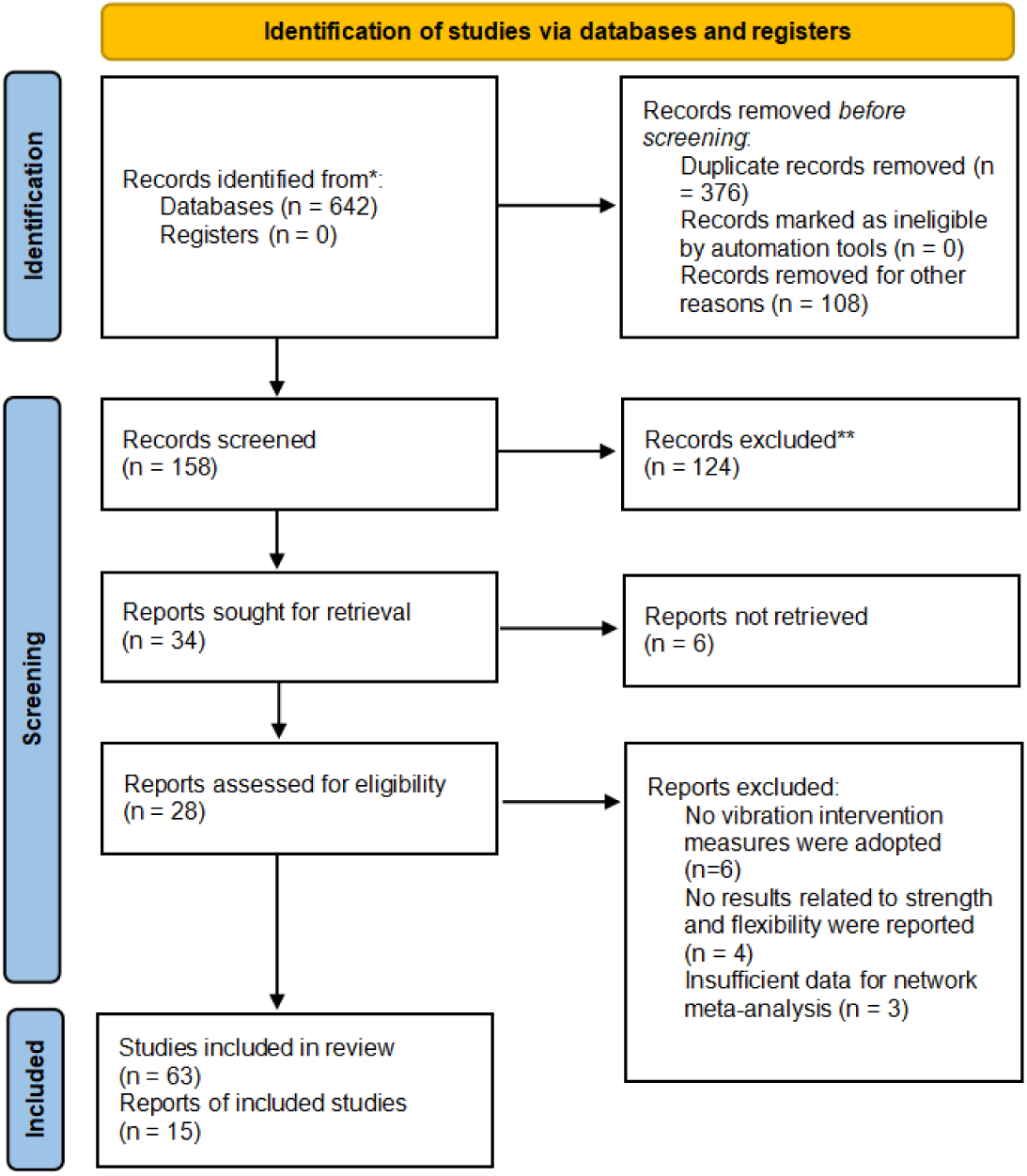
Flowchart.

### 3.2 Characteristics of Included Studies

A total of 15 studies comprising 62 independent trials with 491 participants were included in this meta-analysis. The age of participants ranged from 15 to 79 years. In terms of population types, six studies focused on athletic populations (including basketball, speed skating, hockey, and sprinting), while the remaining nine studies investigated non-athletic individuals or special populations (e.g., postmenopausal women, overweight/obese individuals). All included studies established control groups to compare the effects of whole-body vibration (WBV) training against conventional training or no intervention. Regarding outcome measures, 14 studies evaluated muscle strength as the primary indicator, and six studies reported changes in flexibility either exclusively or concurrently. The detailed characteristics of the included studies are presented in Table 1.

**Table 1.**
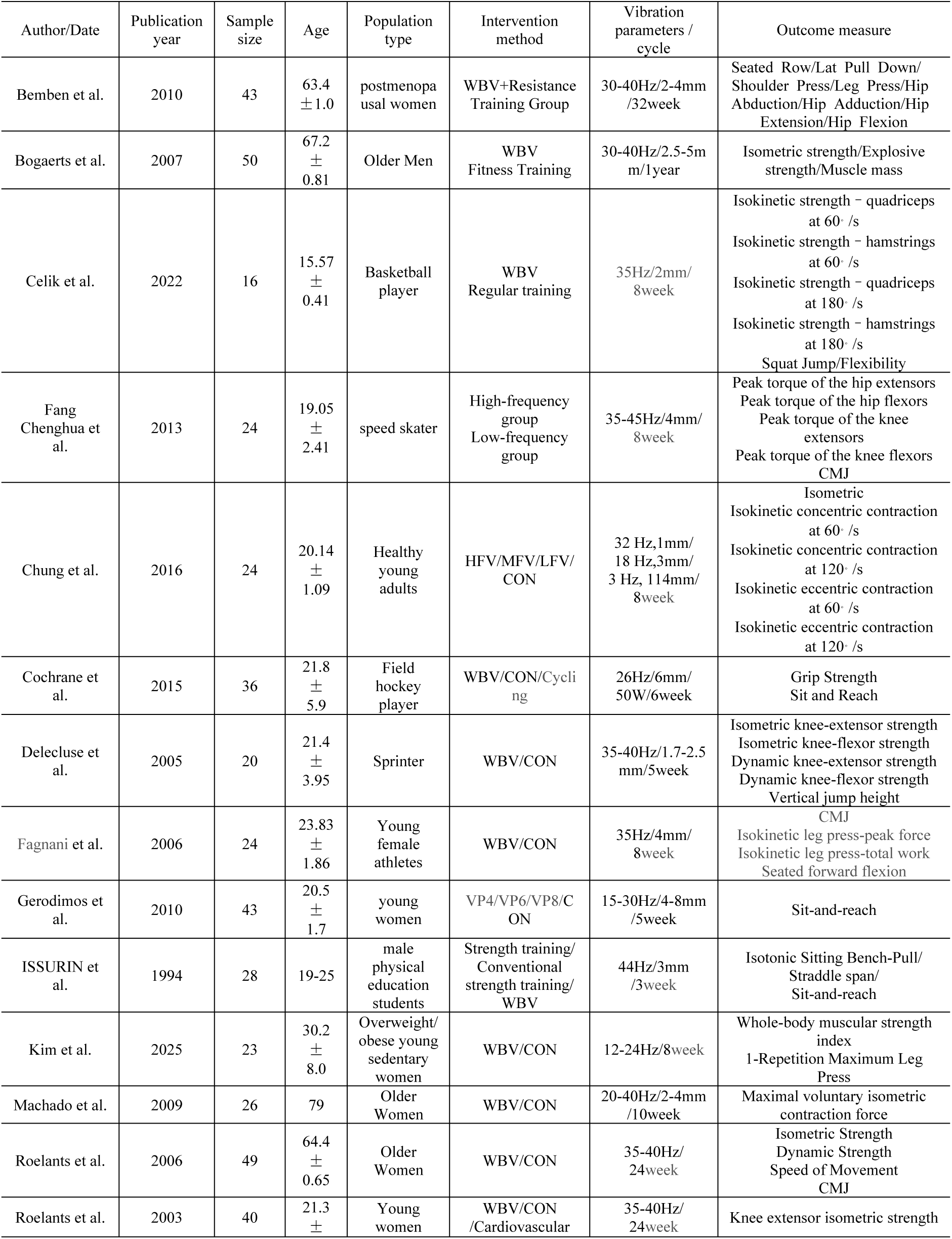
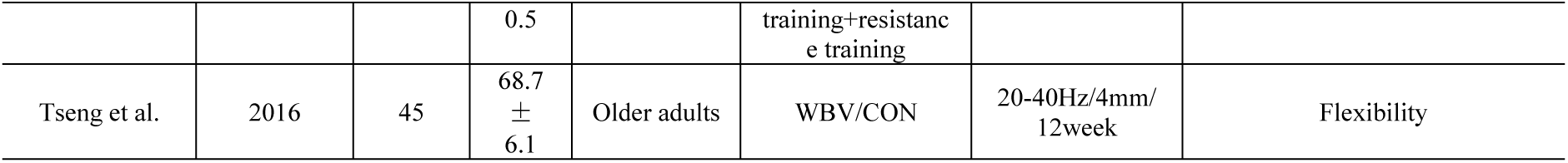
Inclusion of literature information.

### 3.3 Quality Assessment of Included Studies

Quality assessment was conducted on the 62 included studies. Among these, 15 studies described their randomization methods, one study had an unclear risk of bias regarding randomization, and four studies utilized non-randomized controlled designs. The results are shown in Figures 2 and 3.

**Figure 2.**
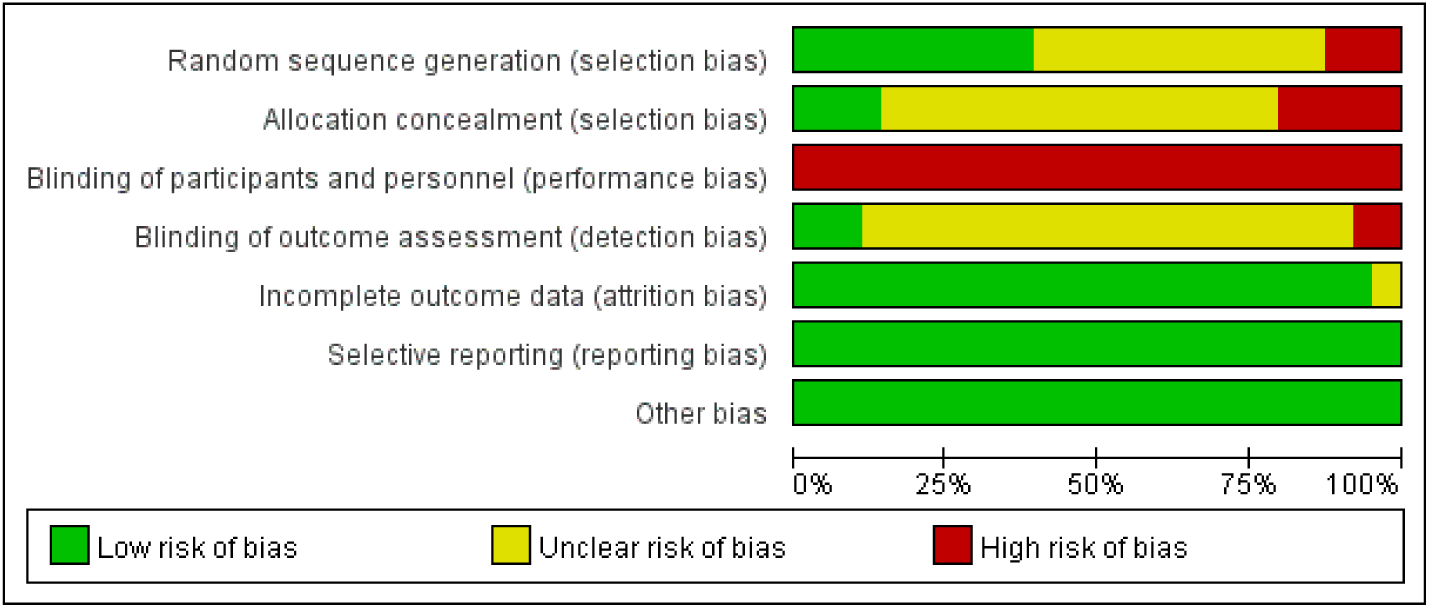
Percentage of each item in the methodological quality assessment.

**Figure 3.**
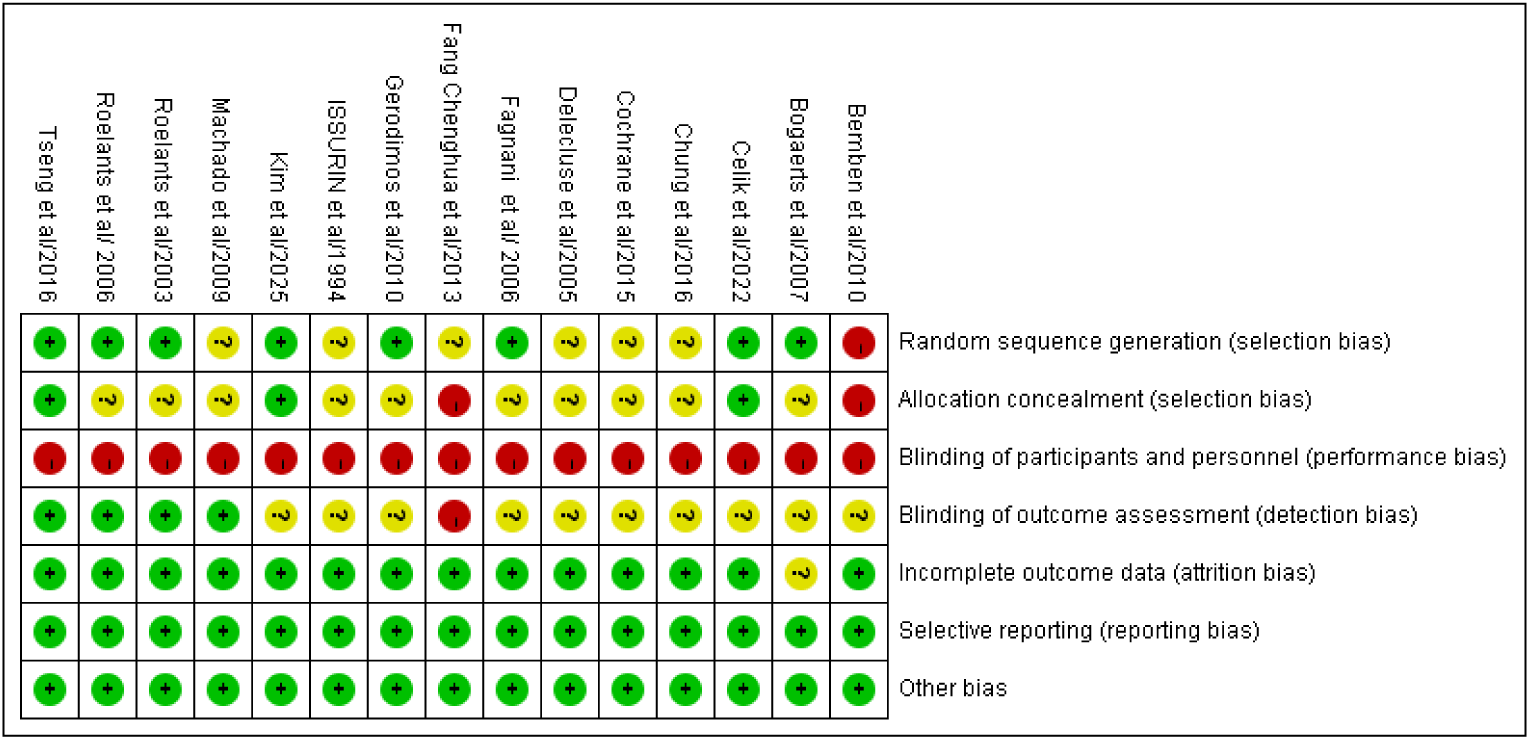
Percentage of each item in the methodological quality assessment.

### 3.4 Meta-Analysis Results

To evaluate the influence of individual studies on the pooled effect size and the stability of the results, a leave-one-out sensitivity analysis was performed. Specifically, each included study was sequentially removed, and the pooled effect size (Standardized Mean Difference, SMD) with its 95% confidence interval (CI) was recalculated for the remaining studies. The fluctuations in the pooled effect size before and after exclusion were visually compared and presented in Figure 4.

**Figure 4.**
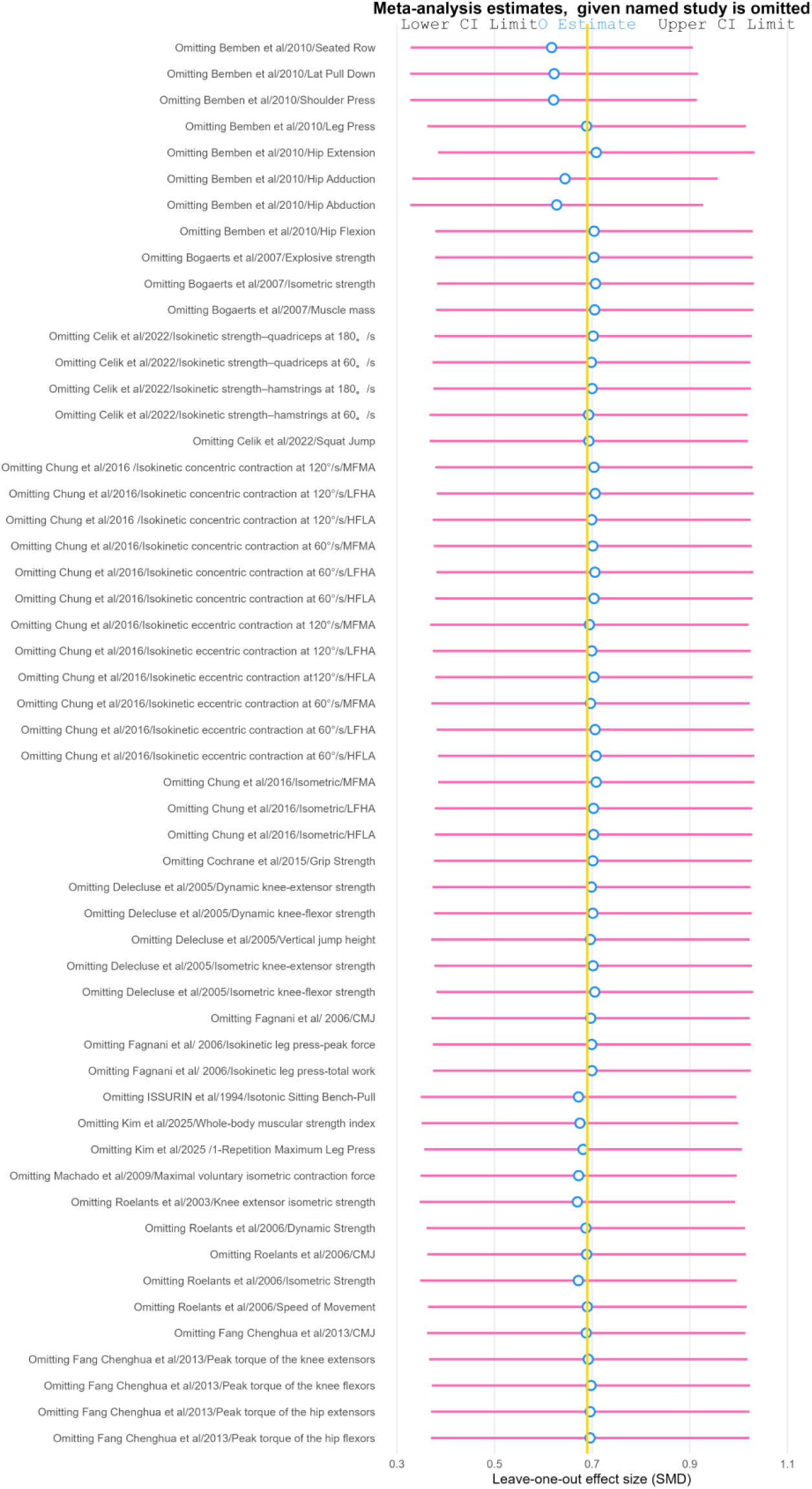
Leave-one-out sensitivity analysis.

The results demonstrated that after sequentially removing each included study, the estimated pooled effect sizes (blue dots) all fell within the 95% CI range of the original pooled effect size. Furthermore, the 95% CIs calculated after excluding any single study did not cross the line of null effect (SMD = 0). This indicates that no individual study exerted a dominant influence on the overall pooled results, confirming the robustness and reliability of the findings in this meta-analysis.

#### Flexibility

For the flexibility intervention subgroup, a total of 8 studies were included, comprising a total of 242 participants. The forest plot is shown in Figure 5.

**Figure 5.**
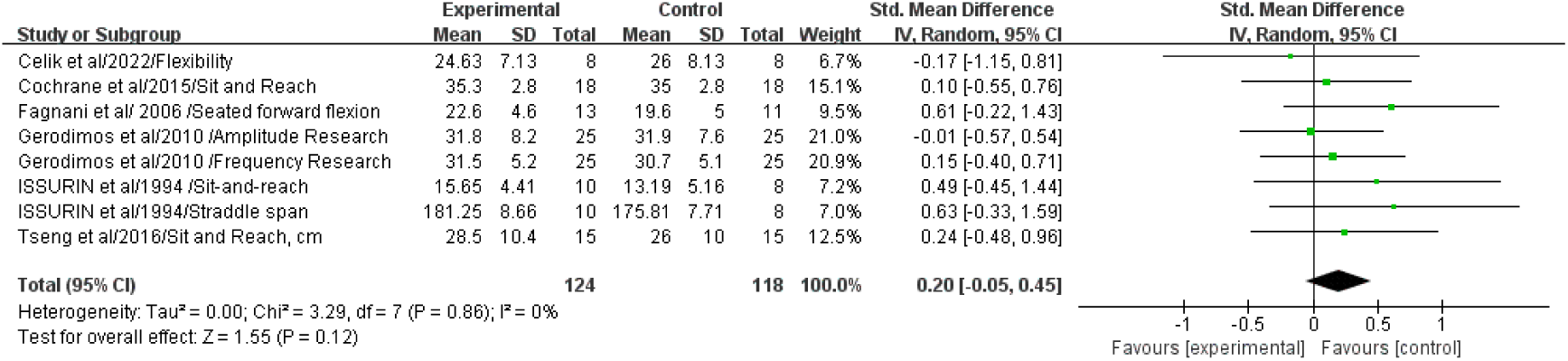
Forest plot of the flexibility intervention subgroup.

As shown in Figure 5, the heterogeneity among studies in this subgroup was negligible (P = 0.86, I ² = 0%). A fixed-effects model was adopted for analysis, yielding a pooled effect size of SMD = 0.20, 95% CI [-0.05, 0.45], indicating a small and non-significant effect of vibration training on flexibility improvement.

#### Strength

##### Subgroup Analysis by Population

As shown in Figure 6,in the elderly subgroup, a total of 8 studies involving 372 participants were included. Meta-analysis results indicated considerable heterogeneity among the studies in this subgroup (P < 0.0001, I²= 77%). A random-effects model was applied for analysis, yielding a pooled effect size of SMD = 0.65, 95% CI [0.20, 1.10], demonstrating a moderate effect of vibration training on improving strength in the elderly population.

**Figure 6.**
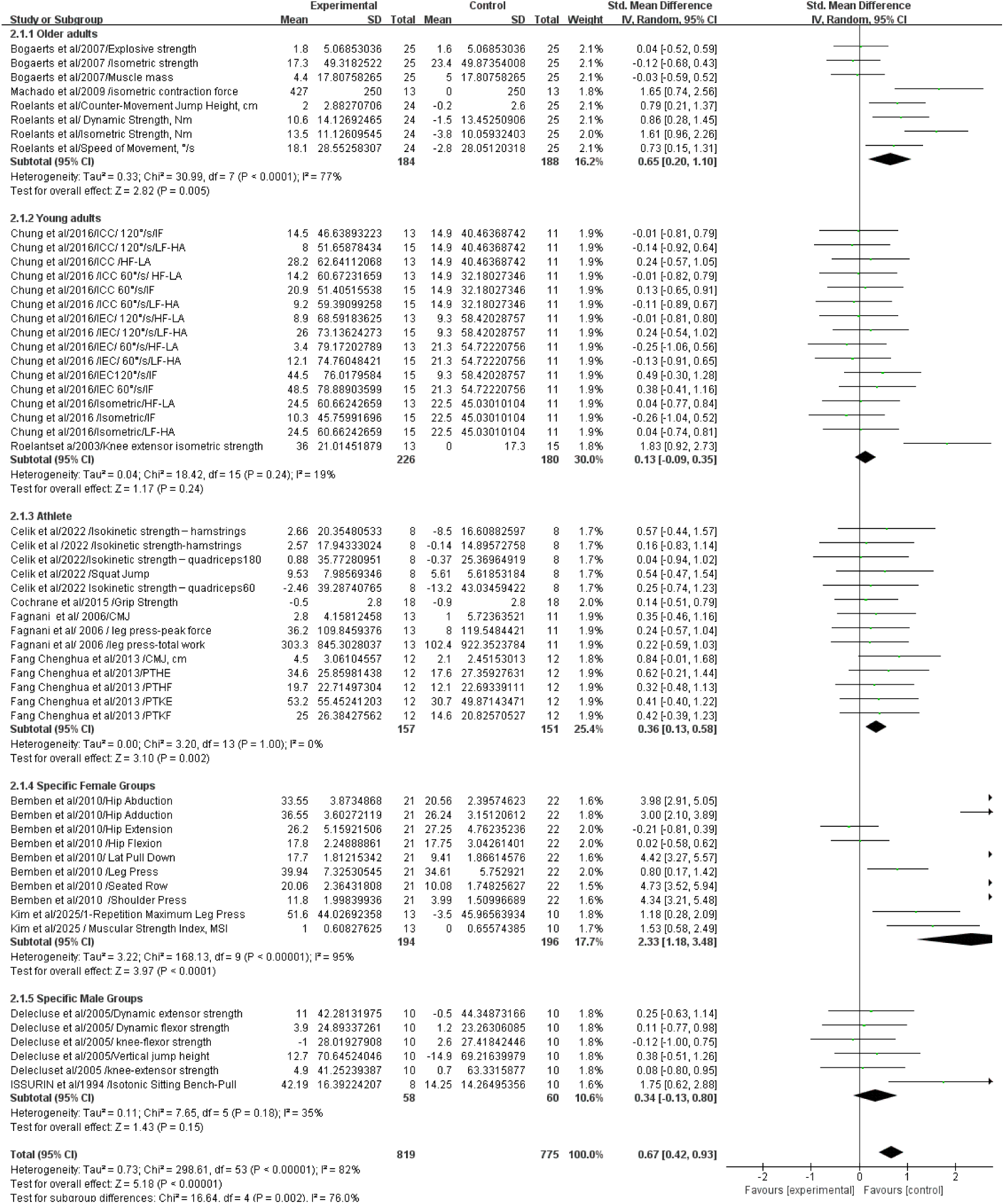
Forest Plot of Subgroup Analyses by Population.

**Figure 7.**
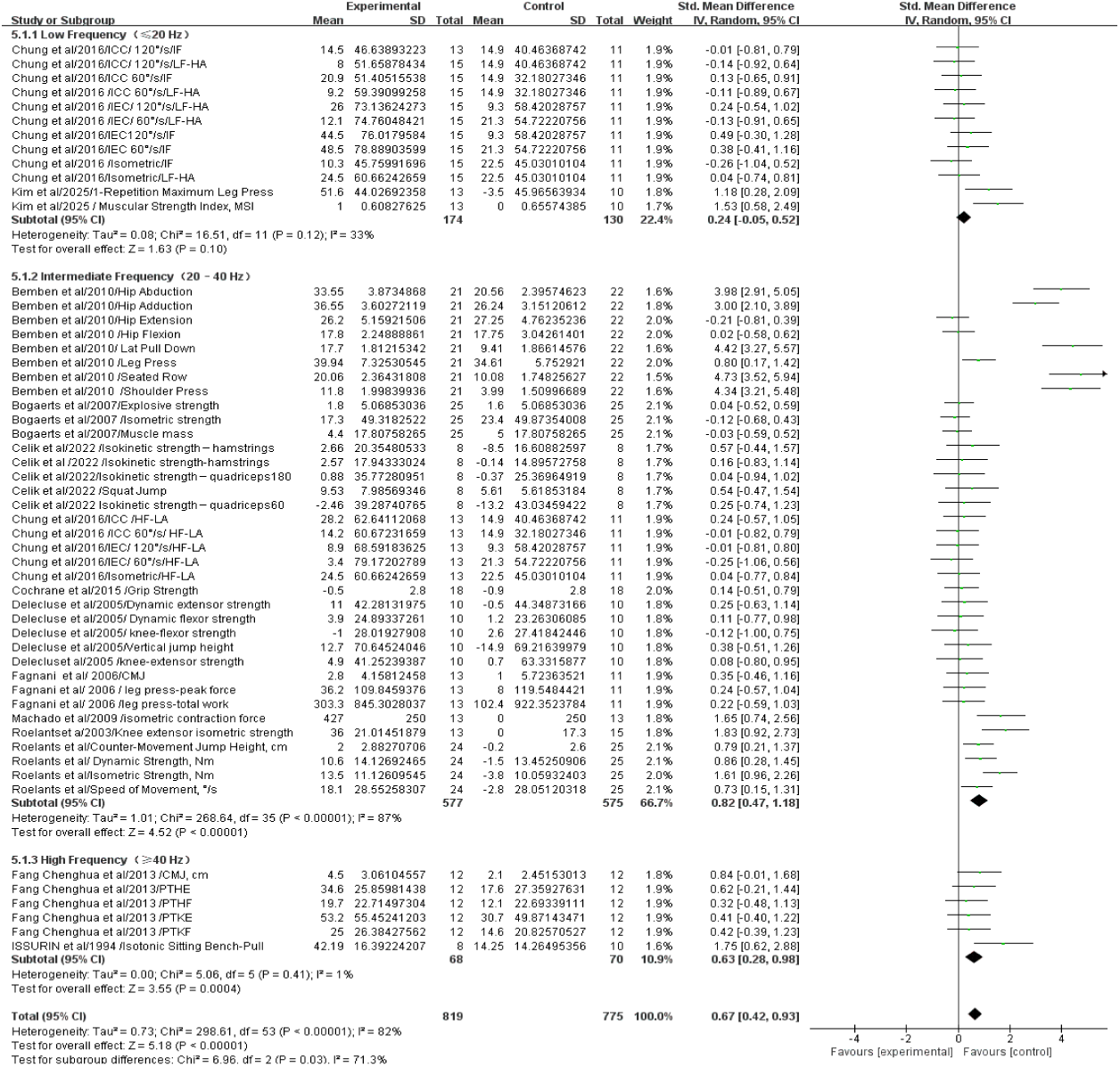
Forest Plot of Subgroup Analysis by Vibration Frequency

**Figure 8.**
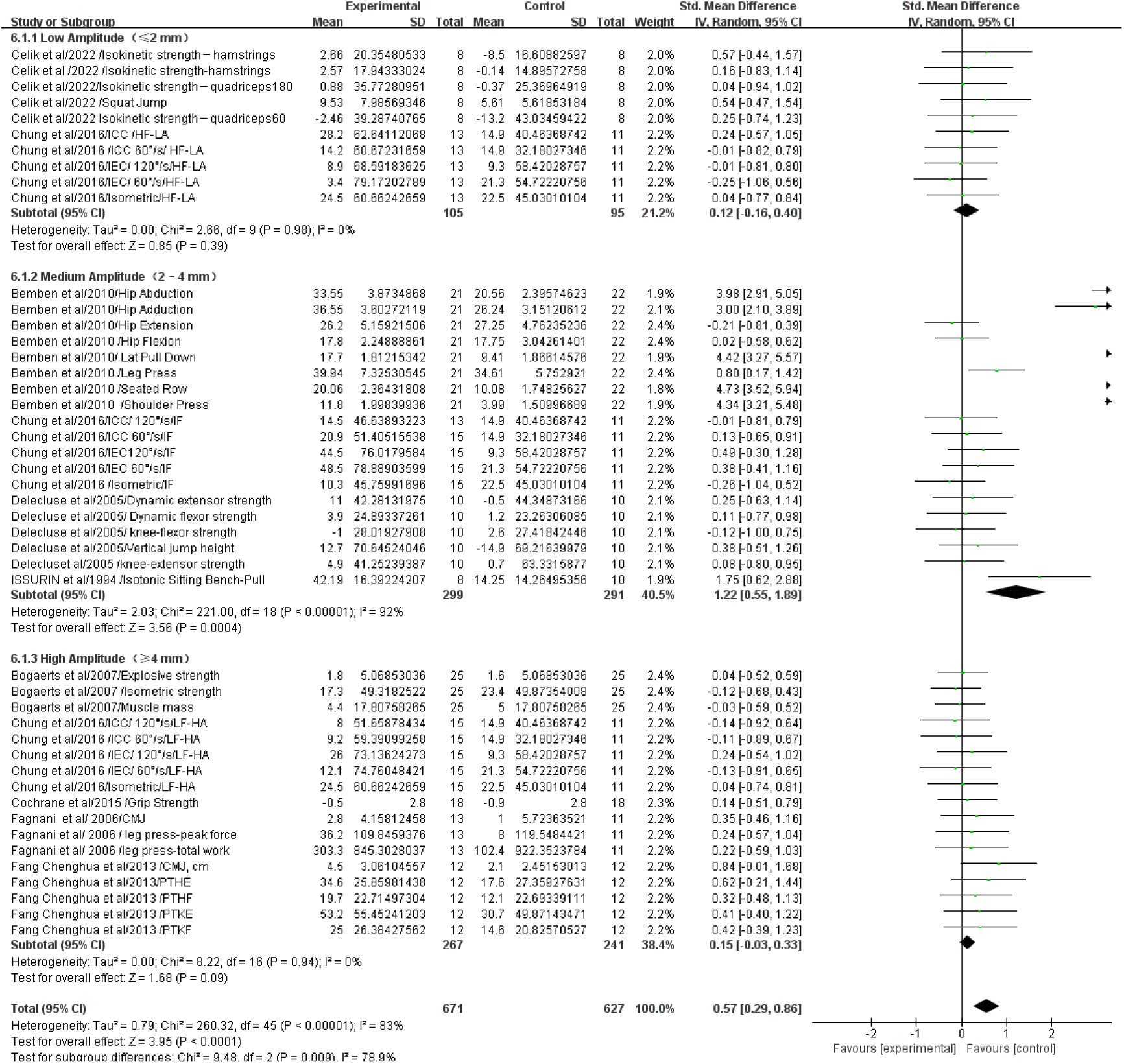
Forest Plot of Subgroup Analysis by Amplitude

**Figure 9.**
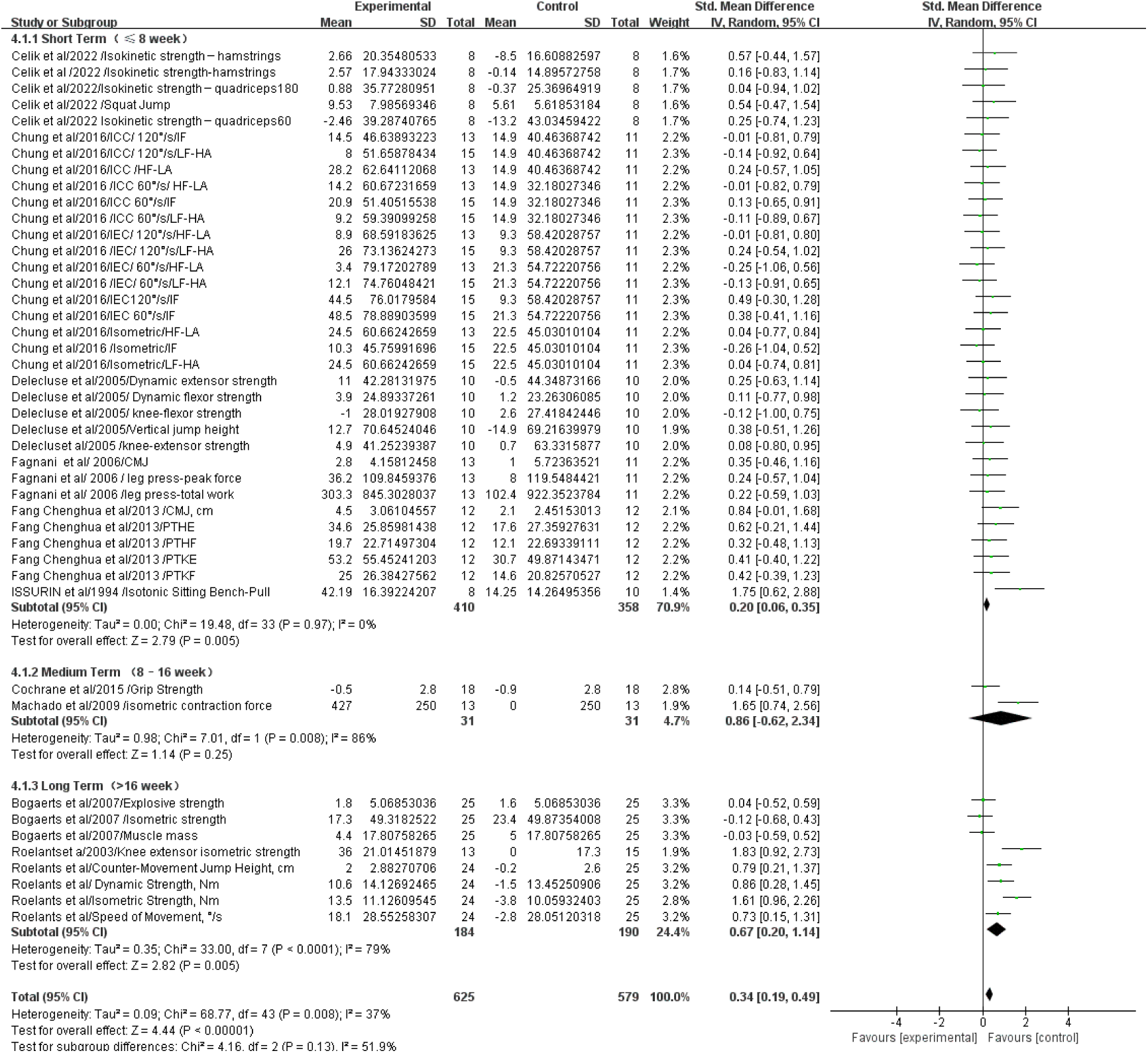
Forest Plot of Subgroup Analysis by Training Duration

In the young adults subgroup, a total of 16 studies involving 406 participants were included. Meta-analysis results indicated low heterogeneity among the studies in this subgroup (P = 0.24, I²= 19%). A fixed-effects model was applied for analysis, yielding a pooled effect size of SMD = 0.13, 95% CI [-0.09, 0.35], indicating a small and non-significant effect of vibration training on strength improvement in young adults.

In the athletes subgroup, a total of 14 studies involving 308 participants were included. Meta-analysis results indicated negligible heterogeneity among the studies in this subgroup (P = 1.00, I²= 0%). A fixed-effects model was applied for analysis, yielding a pooled effect size of SMD = 0.36, 95% CI [0.13, 0.58], indicating a small but statistically significant effect of vibration training on strength improvement in athletes.

In the special female population subgroup, a total of 7 studies involving 390 participants were included. The meta-analysis results demonstrated substantial heterogeneity among these studies (P < 0.0001, I²= 95%). A random-effects model was applied for analysis, yielding a pooled effect size of SMD = 2.33, 95% CI [1.18, 3.48], indicating a large effect of vibration training on strength improvement in special female populations.In the special male population subgroup, a total of 6 studies involving 118 participants were included. Meta-analysis results indicated low heterogeneity among the studies in this subgroup (P = 0.18, I²= 35%). A fixed-effects model was applied for analysis, yielding a pooled effect size of SMD = 0.34, 95% CI [-0.13, 0.80], demonstrating a moderate yet statistically non-significant effect of vibration training on strength improvement in special male populations.

##### Subgroup Analysis by Vibration Frequency

In the low-frequency (≤ 20 Hz) subgroup, a total of 12 studies involving 304 participants were included. Meta-analysis results indicated low heterogeneity among the studies in this subgroup (P = 0.12, I² = 33%). A fixed-effects model was applied for analysis, yielding a pooled effect size of SMD = 0.24, 95% CI [-0.05, 0.52], suggesting a small and non-significant effect of low-frequency vibration on strength improvement.In the medium-frequency (20-40 Hz) subgroup, a total of 36 studies involving 1,152 participants were included. Meta-analysis results revealed substantial heterogeneity among these studies (P < 0.00001, I² = 87%). A random-effects model was consequently applied, yielding a pooled effect size of SMD = 0.82, 95% CI [0.47, 1.18], indicating a large effect of medium-frequency vibration on strength improvement.In the high-frequency (≥40 Hz) subgroup, a total of 6 studies involving 138 participants were included. Meta-analysis results indicated negligible heterogeneity among the studies in this subgroup (P = 0.41, I² = 1%). A fixed-effects model was applied for analysis, yielding a pooled effect size of SMD = 0.63, 95% CI [0.28, 0.98], demonstrating a moderate effect of high-frequency vibration on strength improvement.

##### Subgroup Analysis by Amplitude

In the low-amplitude (≤ 2 mm) subgroup, a total of 10 studies involving 200 participants were included. Meta-analysis results indicated negligible heterogeneity among the studies in this subgroup (P = 0.98, I²= 0%). A fixed-effects model was applied for analysis, yielding a pooled effect size of SMD = 0.12, 95% CI [-0.16, 0.40], suggesting a small and non-significant effect of low-amplitude vibration on strength improvement.

In the medium-amplitude (2-4 mm) subgroup, a total of 19 studies involving 590 participants were included. Meta-analysis results revealed substantial heterogeneity among these studies (P < 0.00001,I^2^=92%). A random-effects model was consequently applied, yielding a pooled effect size of SMD = 1.22, 95% CI [0.55, 1.89], indicating a large effect of medium-amplitude vibration on strength improvement.

In the high-amplitude (≥ 4 mm) subgroup, a total of 17 studies involving 508 participants were included. Meta-analysis results indicated negligible heterogeneity among the studies in this subgroup (P = 0.94, I²= 0%). A fixed-effects model was applied for analysis, yielding a pooled effect size of SMD = 0.15, 95% CI [-0.03, 0.33], indicating a small and non-significant effect of high-amplitude vibration on strength improvement.

##### Subgroup Analysis by Training Duration

In the short-term (≤ 8 weeks) subgroup, a total of 34 studies involving 768 participants were included. Meta-analysis results indicated negligible heterogeneity among the studies in this subgroup (P = 0.97, I² = 0%). A fixed-effects model was applied for analysis, yielding a pooled effect size of SMD = 0.20, 95% CI [0.06, 0.35], indicating a small but statistically significant effect of short-term vibration training on strength improvement.

In the medium-term (8-16 weeks) subgroup, a total of 2 studies involving 62 participants were included. Meta-analysis results revealed substantial heterogeneity between these studies (P = 0.008, I ² = 86%). A random-effects model was consequently applied for analysis, yielding a pooled effect size of SMD = 0.86, 95% CI [-0.62, 2.34], indicating a large effect of medium-term vibration training on strength improvement, though the result did not reach statistical significance.

In the long-term (>16 weeks) subgroup, a total of 8 studies involving 374 participants were included. Meta-analysis results indicated considerable heterogeneity among these studies (P = 0.0001, I² = 79%). A random-effects model was applied for analysis, yielding a pooled effect size of SMD = 0.67, 95% CI [0.20, 1.14], demonstrating a moderate effect of long-term vibration training on strength improvement.

### 3.5 Meta-Regression Analysis Univariable Meta-Regression Analysis

Under the random-effects model (REML method), univariable mixed-effects meta-regression analyses were performed on 54 effect sizes, with frequency (freq), amplitude (amp), and cycle (cyc) included separately as moderators. The results indicated a positive, albeit non-significant, trend for frequency (freq) on the effect size (β = 0.0250, SE = 0.0134, z = 1.88, p = 0.0608). Amplitude showed a negative, non-significant trend (β= -0.0069, SE = 0.0047, z = -1.48, p = 0.1390). In contrast, cycle demonstrated a significant positive relationship with the effect size (β= 0.0289, SE = 0.0113, z = 2.56, p = 0.0103), with a 95% confidence interval of [0.0068, 0.0510], indicating that a longer intervention duration significantly predicted greater effect sizes. All models exhibited high residual heterogeneity (I²=86-87%). However, the cycle model yielded the lowest AIC (164.95) and the highest proportion of explained heterogeneity (R²= 8.75%), suggesting it provided the best fit among the univariable models.

#### Multivariable Model

Based on the univariable results, both frequency and cycle were included in a multivariable mixed-effects meta-regression model. The results indicated that the overall moderating effect was statistically significant (QM(2) = 9.08, p = 0.0107). The proportion of explained heterogeneity increased to 11.72% (R ² = 11.72%), and the AIC decreased to 162.62, suggesting a modest improvement in model fit.Within this model, cycle remained a significant positive predictor (β= 0.0261, SE = 0.0112, z = 2.32, p = 0.0205), whereas frequency was no longer significant (β= 0.0202, SE = 0.0131, p = 0.1233). The test for residual heterogeneity remained significant (QE(51) = 283.24, p < .0001), indicating that substantial sources of heterogeneity remained unaccounted for.

#### Final Model

Based on the results of the univariable and multivariable analyses, the final model retained only the significant variable, cycle, as a moderator (Figure 10). The results confirmed that the effect of cycle on the effect size remained significant (β= 0.0289, SE = 0.0113, z = 2.56, p = 0.0103). The model explained approximately 8.8% of the between-study heterogeneity (R²= 8.75%). Nevertheless, residual heterogeneity remained substantial (I ² = 86.23%, QE(52) = 300.19, p < .0001), suggesting the potential presence of other moderating factors beyond cycle duration that were not included in the model.

**Figure 10.**
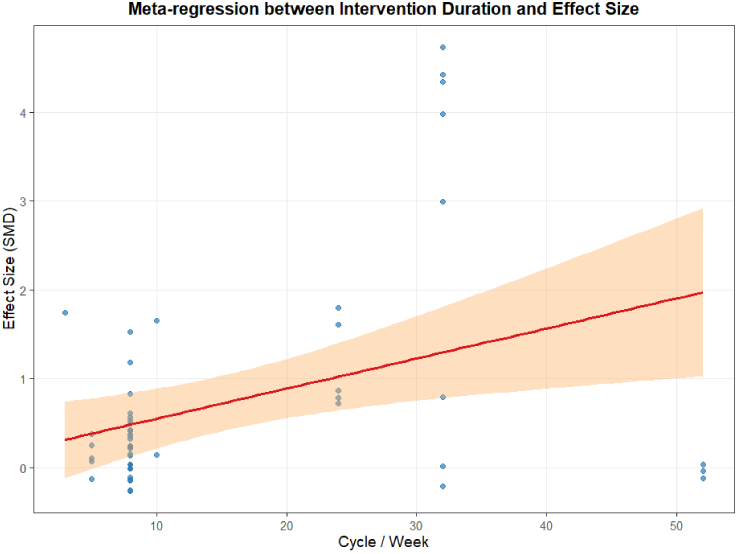
Meta-Regression Plot of Intervention Cycle and Effect Size.

### 3.6 Publication Bias

To assess the potential for publication bias, a funnel plot was initially generated and Egger’s regression test was performed. The results of Egger’s test indicated statistically significant asymmetry in the funnel plot (z = 6.23, p < 0.001), suggesting that smaller studies might be more likely to report larger positive effects. Consequently, Duval & Tweedie’s Trim-and-Fill analysis was further employed. This analysis imputed zero missing studies (Estimated Missing Studies = 0, SE = 2.80). The adjusted pooled effect size after trim-and-fill (g = 0.56, 95% CI [0.35, 0.76]) was identical to the original random-effects model estimate. In other words, the Trim-and-Fill method did not indicate a substantial impact of publication bias on the overall effect, as shown in Figure 11.

**Figure 11.**
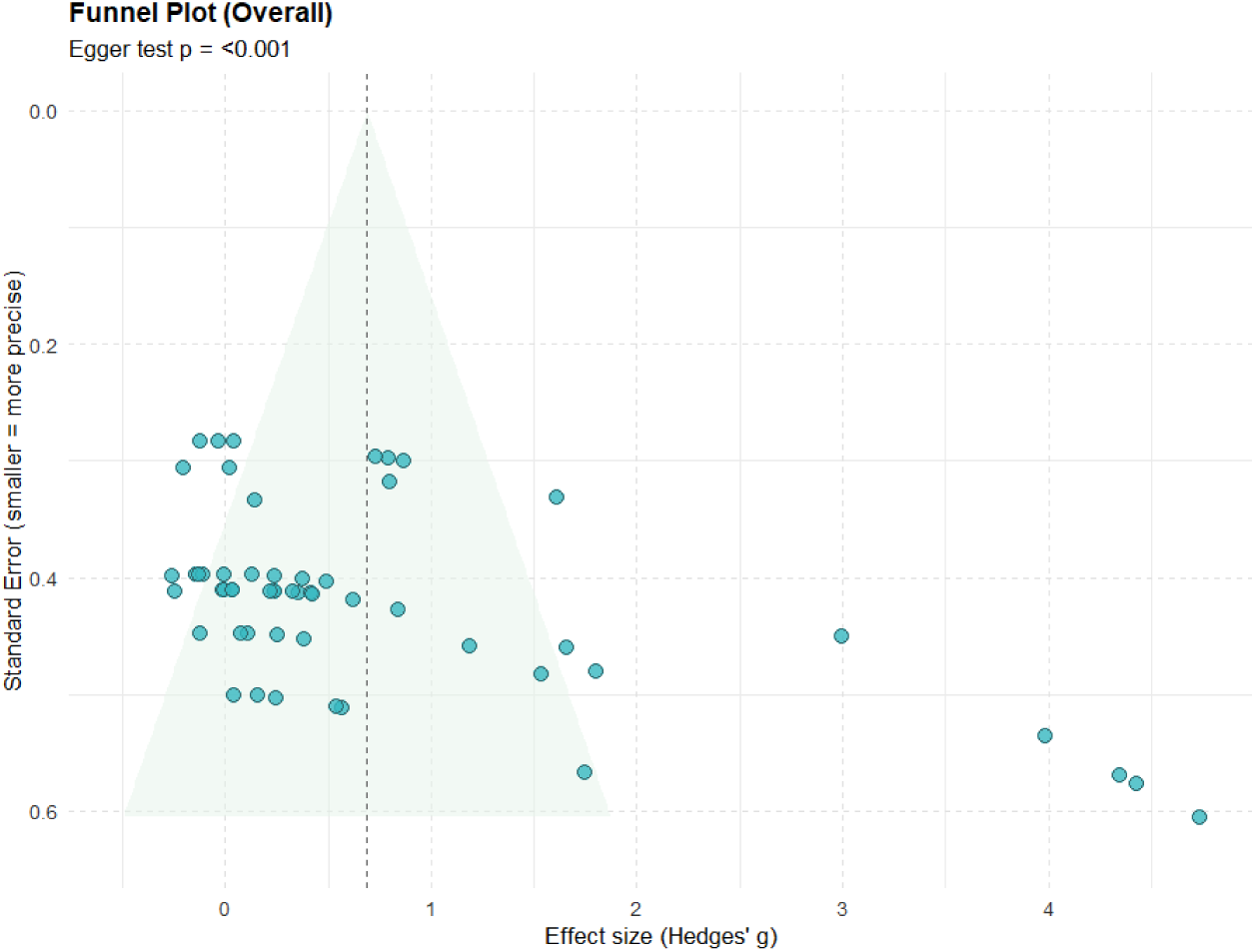
Trim-and-Fill Funnel Plot for Publication Bias Analysis.

Furthermore, the high heterogeneity observed in this study (I²= 82%) indicates substantial methodological or sample differences among the included studies. Elevated heterogeneity can potentially lead to false-positive signals in Egger’s test. Therefore, based on a comprehensive assessment, although Egger’s test indicated funnel plot asymmetry, both the trim-and-fill analysis and effect robustness tests did not reveal substantial publication bias. Collectively, the findings of this study are robust, and the pooled effect size (g≈0.56) demonstrates a moderate level of practical significance.

## 4 Discussion

This meta-analysis demonstrates that whole-body vibration training has a significant effect on strength capacity, with WBV demonstrating a broadly positive effect on muscle strength enhancement. The underlying physiological mechanism aligns with the findings of Hagbarth & Eklund, indicating that vibration stimuli activate the primary endings of muscle spindles (Group Ia afferent fibers), thereby potentiating the stretch reflex arc and eliciting involuntary muscle contractions, known as the Tonic Vibration Reflex (TVR) (Hagbarth & Eklund, 1966). This neuro-driven contraction pattern efficiently recruits motor units, particularly fast-twitch fibers, and optimizes motor unit synchronization. Consequently, it enhances the excitability and power output of the neuromuscular system without requiring significant external load (Cardinale & Bosco, 2003). This is especially relevant for populations who cannot tolerate high-load resistance training, such as older adults and rehabilitation patients.The subgroup analysis in this study clearly reveals substantial differences in responses to vibration stimuli across populations. In the elderly subgroup, WBV demonstrated a moderate and significant effect (SMD = 0.65). This is closely related to the prevalent context of sarcopenia and neuromuscular function decline in older adults (Cruz-Jentoft et al., 2019). With aging, the degeneration of alpha motor neurons, decreased muscle spindle sensitivity, and hormonal changes collectively contribute to the loss of muscle mass and strength (Delmonico et al., 2009). While traditional resistance training has proven effective, issues related to adherence, safety, and accessibility remain (Chodzko et al., 2009). WBV, as a low-load, high-frequency neuromuscular stimulus, effectively addresses this gap. It may work by reactivating “dormant” motor neurons and improving neural drive efficiency, thereby facilitating strength gains even before noticeable muscle hypertrophy occurs (Machado et al., 2010).

In contrast, athletes and healthy young adults, who are already in a state of high neuromuscular adaptation due to the considerable intensity, volume, and specificity of their regular training, exhibited only small effect sizes from WBV interventions. The effects were minimal in young adults (SMD = 0.13) and small in athletes (SMD = 0.36), with the improvement in strength among general young adults being non-significant.The additional stimulus provided by WBV may fail to surpass the activation threshold of their neuromuscular systems, thus becoming submerged within the potent effects of their foundational training—a phenomenon known as the “ceiling effect” (Cormie, McBride, & McCaulley, 2007). Furthermore, athletic training is highly sport-specific. If the postures or movement patterns adopted during WBV are not closely aligned with specific technical actions, the resulting neural adaptations may not effectively transfer to sports performance (Young, 2006). Therefore, for high-performance populations, the application of WBV requires more refined strategies. It should be integrated into specific phases such as warm-up activation, recovery, and regeneration, or used to address targeted weaknesses, rather than serving as a direct substitute for core strength training.

The study further confirmed that the special female population subgroup exhibited a very large effect size (SMD = 2.33), which was accompanied by extremely high heterogeneity (I ² = 95%). This represents a highly promising yet complex area requiring clarification. The substantial effect may stem from this group’s unique physiological responsiveness to vibration stimuli. This subgroup likely comprises diverse subpopulations, including postmenopausal women, young female athletes, and postpartum women, each with distinct physiological states, hormonal profiles, and training backgrounds, leading to varied responses to vibration (Janse de Jonge, 2003). For instance, postmenopausal women, due to a sharp decline in estrogen levels, represent a high-risk group for sarcopenia and osteoporosis (Greising, Baltgalvis, & Lowe, 2009).

The efficacy of WBV depends not only on “for whom it is used” but also critically on “how it is applied.” The parameter-based subgroup analysis in this study provides crucial insights for precise modulation in practice. The findings indicate that medium-frequency (20–40 Hz, SMD = 0.82) and high-frequency (≥40 Hz, SMD = 0.63) vibrations yield superior strength improvements compared to low-frequency (≤ 20 Hz, SMD = 0.24) vibrations. From a physiological perspective, muscle spindles are most sensitive to vibratory stimuli within the 20–50 Hz range. Within this frequency band, Group Ia afferent fibers can generate high-frequency, sustained discharges, thereby maximally activating spinal alpha motor neurons and eliciting potent muscle contractions (Schieppati, 1987).Excessively low vibration frequencies (e.g., <20 Hz) may be insufficient to induce tetanic contractions, while excessively high frequencies (e.g., >50 Hz) could reduce contraction efficiency due to overly rapid firing rates and may even trigger protective inhibition. It is noteworthy that the medium-frequency subgroup exhibited extremely high heterogeneity (I²= 87%), indicating that the pursuit of an “ optimal f requency” in i solation i s one-sided—frequency must be considered in conjunction with other parameters (particularly amplitude) and individual perception. Amplitude, which determines the intensity of the vibration stimulus, is another core mechanical parameter influencing training effects(Liu et al., 2009). This study identified a clear and significant “medium-amplitude effect” (2-4 mm): this amplitude range not only yielded the largest effect size (SMD = 1.22, large effect) but also significantly outperformed both the low-amplitude (≤2 mm) and high-amplitude (≥4 mm) subgroups.Excessively low amplitudes may fail to provide sufficient mechanical stimulation to effectively activate muscle spindles, whereas excessively high amplitudes may induce excessive body oscillation. To maintain postural stability, the body may activate antagonistic muscle co-contraction, thereby reducing the net force output of the target muscle groups and simultaneously compromising training comfort and compliance. Medium amplitude, in contrast, strikes an optimal balance between stimulation intensity and bodily controllability, thereby optimizing neuromuscular response efficiency. Training duration reflects the cumulative effect of training stimuli, and the results clearly demonstrate the temporal pattern of training adaptation. Short-term training (≤ 8 weeks) primarily induces neural adaptations, such as improvements in motor unit recruitment and firing rates, resulting in a small effect size (SMD = 0.20). In contrast, medium-term and long-term training (8-16 weeks, >16 weeks) yielded large and moderate effect sizes (SMD = 0.86 and SMD = 0.67, respectively), suggesting that sustained WBV stimulation may be sufficient to induce long-term structural and functional muscular adaptations, such as muscle fiber hypertrophy (Machado et al., 2010). This aligns with the fundamental physiological principle of training adaptation: neural adaptations occur rapidly and early, while structural adaptations develop more slowly and require longer durations (Folland & Williams, 2007). Therefore, to achieve substantial and sustained strength gains through WBV, a sufficiently long intervention period is essential, positioning it as a medium- to long-term training strategy rather than a short-term solution.

In contrast to its effects on strength, the impact of WBV on flexibility appears uncertain and yields only a small effect size (SMD = 0.20). This may be attributed to its inherent mechanistic limitations. Improvements in flexibility primarily depend on the elongation of the muscle-tendon unit and an increase in joint range of motion, which are typically achieved through sustained stretching (Weppler & Magnusson, 2010). Although WBV may induce repeated muscle contractions and relaxations via the TVR and potentially produce transient improvements in viscoelastic properties by raising muscle temperature and enhancing blood flow, these effects are likely brief and unstable, failing to induce long-term morphological adaptations in connective tissues. Furthermore, the limited number of studies in this area (only 8) results in low statistical power, contributing to the high uncertainty of the current conclusions.

### Limitations of This Study

Substantial variations in vibration equipment models, participant positioning, and combined training protocols among the included studies were the primary sources of heterogeneity. The limited number of studies in certain subgroups may have affected the stability of the results. Furthermore, incomplete or imprecise reporting of vibration parameters (e.g., amplitude) in many original studies posed challenges for accurate data extraction and subgroup analysis.

### Future Research Directions

1)Conduct high-quality, large-sample randomized controlled trials (RCTs), with particular focus on special populations such as women and individuals with chronic diseases, and implement rigorous subgroup stratification.2)Deepen mechanistic investigations by integrating advanced techniques such as surface electromyography (sEMG), mechanomyography (MMG), blood biomarkers, and muscle imaging (e.g., ultrasonography, MRI) to elucidate the effects of WBV from multiple dimensions, including neural, endocrine, and muscular structural adaptations.3)Emphasize individualization and specificity by examining how individual differences (e.g., training background, genotype, physique) influence responses to vibration, and develop vibration training protocols closely aligned with specific sports disciplines. Amplitude should be regarded as a core parameter equally important as frequency, and blind pursuit of high amplitude should be avoided.4)Future studies should focus on well-designed research to explore whether combining WBV with traditional methods such as dynamic stretching and PNF stretching can produce synergistic effects.

## 5 Conclusion

This study adopted a rigorous meta-analytic approach to synthesize existing evidence on the effects of whole-body vibration training on strength and flexibility. Through an in-depth deconstruction of population characteristics, vibration parameters, and training duration, the following conclusions and practical recommendations are drawn,1)Confirmed Overall Efficacy: Whole-body vibration training, as an auxiliary training modality, demonstrates a moderate positive effect on improving muscle strength (SMD = 0.67). However, its benefits for flexibility remain inconclusive based on current evidence.2)Pronounced Population-Specific Effects: Older adults are among the primary beneficiaries of WBV, with moderate effect sizes confirming its role as an effective strategy against age-related strength decline. The limited effects observed in athletes and healthy young adults suggest WBV should be positioned as a supplementary, rather than core, training method for these groups. Special female populations show significant potential, although the highly unstable effects highlight a critical need for future targeted research.3)Existence of an “Optimal Window” for Vibration Parameters:1)Frequency:Medium to high frequencies(≥ 20Hz) are superior to low frequencies for strength enhancement.2)Amplitude: Medium amplitude (2-4 mm) is crucial for optimal strength training outcomes, proving significantly more effective than both low and high amplitudes, thereby dispelling the misconception that “larger amplitude is always better.3)Training Duration: Medium- to long-term interventions yield superior results compared to short-term programs, confirming WBV’s potential for inducing long-term structural adaptations.

Based on the above conclusions, specific recommendations are provided for practitioners in different fields:1)For Rehabilitation and Geriatrics: It is strongly recommended to incorporate WBV into the regular physical training plans for older adults, post-operative rehabilitation patients, and individuals with chronic conditions. Starting parameters are suggested at medium frequency (25-35 Hz) and medium amplitude (2-3 mm), with a frequency of 2-3 sessions per week, maintained long-term (for at least 12–16 weeks). This aims to improve neuromuscular function, delay muscle atrophy, and reduce fall risk.2)For Competitive Sports: Coaches should adopt a rational perspective on the role of WBV. It can be utilized during the warm-up and activation phase. Employ medium-to-high frequency and low-to-medium amplitude vibrations pre-competition or pre-training to rapidly enhance neural excitability and core temperature. During the recovery and regeneration phase, use low-frequency, low-amplitude vibrations post-training to promote blood circulation and alleviate muscle stiffness. For strengthening weak links, apply targeted stimulation to small or hard-to-activate muscle groups. It is crucial to avoid expecting WBV to completely replace traditional resistance and sport-specific strength training.3)For Public Fitness and Special Populations: WBV offers a safe and effective entry-level option for physically weak individuals, beginners, or those with joint load limitations. For female populations, particularly perimenopausal and postmenopausal women, WBV demonstrates excellent application potential. However, personalized program design under the guidance of professionals is recommended.

In conclusion, whole-body vibration training is by no means a “universal” training solution. Its value lies in its scientific and individualized application. The findings of this study provide a robust evidence base and a clear practical pathway for understanding “why,” “for whom,” and “how” to utilize WBV across different contexts.

## Competing Interests

The authors declare no competing interests.

The data that support the findings of this study are available from the corresponding author upon reasonable request.

## Data Availability

All relevant data are within the manuscript and its Supporting Information files

